# Investigating associations of omega-3 fatty acids, lung function decline, and airway obstruction

**DOI:** 10.1101/2023.01.18.23284671

**Authors:** Bonnie K. Patchen, Palavi Balte, Traci M. Bartz, R. Graham Barr, Myriam Fornage, Mariaelisa Graff, David R Jacobs, Ravi Kalhan, Rozenn N. Lemaitre, George O’Connor, Bruce Psaty, Jungkyun Seo, Michael Y. Tsai, Alexis C. Wood, Hanfei Xu, Jingwen Zhang, Sina A. Gharib, Ani Manichaikul, Kari North, Lyn M. Steffen, Josée Dupuis, Elizabeth Oelsner, Dana B. Hancock, Patricia A. Cassano

**Affiliations:** Division of Nutritional Sciences, Cornell University, Ithaca, NY; Department of Medicine, Columbia University College of Physicians and Surgeons, New York, NY; Cardiovascular Health Research Unit, Departments of Medicine, Epidemiology, Health Systems and Population Health, University of Washington, Seattle, WA; Brown Foundation Institute of Molecular Medicine, McGovern Medical School, University of Texas Health Science Center, Houston, TX; Department of Epidemiology, University of North Carolina, Chapel Hill, NC; Department of Laboratory Medicine and Pathology, University of Minnesota, Minneapolis, MN; Northwestern Medicine, Chicago, IL; Boston University School of Medicine, Boston, MA; USDA/ARS Children’s Nutrition Research Center, Houston, TX; Boston University School of Public Health, Boston, MA; Division of Pulmonary, Critical Care and Sleep Medicine, University of Washington, Seattle, WA; Center for Public Health Genomics, University of Virginia, Charlottesville, VA; Division of Epidemiology and Community Health, University of Minnesota School of Public Health, Minneapolis, MN; Department of Epidemiology, Biostatistics and Occupational Health, School of Population and Global Health, McGill University, Montréal, Québec; RTI International, Research Triangle Park, NC; Department of Population Health Sciences, Weill Cornell Medicine, New York, NY

**Keywords:** spirometry, nutrition, epidemiology, Mendelian Randomization

## Abstract

**Rationale:** Inflammation contributes to lung function decline and the development of chronic obstructive pulmonary disease. Omega-3 fatty acids have anti-inflammatory properties and may benefit lung health.

**Objectives:** Investigate associations of omega-3 fatty acids with lung function decline and incident airway obstruction in adults of diverse races/ethnicities from general population cohorts.

**Methods:** Complementary study designs: (1) longitudinal study of plasma phospholipid omega-3 fatty acids and repeated FEV_1_ and FVC measures in the National Heart, Lung, and Blood Institute Pooled Cohorts Study, and (2) two-sample Mendelian Randomization (MR) study of genetically predicted omega-3 fatty acids and lung function parameters.

**Measurements and Main Results:** The longitudinal study found that higher omega-3 fatty acid concentrations were associated with attenuated lung function decline in 15,063 participants, with the largest effect sizes for docosahexaenoic acid (DHA). One standard deviation higher DHA was associated with an attenuation of 1.8 mL/year for FEV_1_ (95% confidence interval [CI] 1.3–2.2) and 2.4 mL/year for FVC (95% CI 1.9–3.0). One standard deviation higher DHA was also associated with a 9% lower incidence of spirometry-defined airway obstruction (95% CI 0.86–0.97). DHA associations persisted across sexes, smoking histories, and Black, white and Hispanic participants, with the largest magnitude associations in former smokers and Hispanics. The MR study showed positive associations of genetically predicted omega-3 fatty acids with FEV_1_ and FVC, with statistically significant findings across multiple MR methods.

**Conclusions:** The longitudinal and MR studies provide evidence supporting beneficial effects of higher circulating omega-3 fatty acids, especially DHA, on lung health.

## INTRODUCTION

Lung function increases through early adulthood, plateaus, and then declines with aging. Accelerated lung function decline increases the risk of chronic obstructive pulmonary disease (COPD), a leading cause of death in the United States. The primary risk factor for both accelerated lung function decline and COPD is cigarette smoking,^1^ which increases inflammation in the lungs. Nutrients like omega-3 fatty acids with anti-inflammatory properties represent biologically plausible agents for slowing the rate of lung function decline and preventing the development of chronic airway obstruction.

The omega-3 fatty acids include alpha-linolenic acid (ALA), eicosapentaenoic acid (EPA), docosapentaenoic acid (DPA), and docosahexaenoic acid (DHA). ALA—present in some vegetable oils—must be obtained through the diet, while EPA, DPA, and DHA—found mainly in fish—can also be derived endogenously from ALA.^2^ Metabolism of EPA, DPA, and DHA contributes to total body anti-inflammatory function. Metabolites of these fatty acids help resolve inflammation by blocking recruitment and inducing clearance of immune cells from sites of inflammation and by downregulating the expression of transcriptional factors like NF-κB that increase expression of inflammatory cytokines.^3, 4^

Cross-sectional and case-control studies have reported that higher circulating omega-3 fatty acids correlate with higher lung function and lower risk of airway impairment.^5-9^ To our knowledge, no studies have investigated the relationship of circulating omega-3 fatty acids with subsequent decline in lung function and development of COPD in generally healthy adult populations, limiting causal inference. Randomized controlled trials of nutrition interventions for chronic disease outcomes like COPD are challenging, as most chronic diseases have long latency periods and the optimal window for interventions is unknown.^10, 11^ Alternative study designs to investigate the role of nutrition in the etiology of COPD are needed.

Longitudinal prospective cohorts and Mendelian Randomization (MR) provide two study designs to test whether observational data are consistent with the expectations of a causal relationship. Longitudinal prospective cohorts measure exposures prior to the occurrence of the outcome, thus mitigating concerns about reverse causality. MR studies, which examine genotypes associated with an exposure for relation to an outcome, help mitigate confounding bias by leveraging the inherent randomization of genotypes. We conducted a longitudinal observational study of circulating omega-3 fatty acids and lung function decline and a two-sample MR study of genetically predicted omega-3 fatty acids and lung function parameters. By triangulating evidence from these two study designs, our goal was to improve causal inference for associations of omega-3 fatty acids with longitudinal decline in lung function.

## METHODS

We applied two complementary study designs: 1) a longitudinal observational study of omega-3 fatty acids and repeated measures of forced expiratory volume in the first second (FEV_1_) and forced vital capacity (FVC) in the NHLBI Pooled Cohorts Study, and 2) a two-sample MR study using publicly available genome-wide association study (GWAS) data of omega-3 fatty acid biomarkers, FEV_1_, FVC, and spirometry-defined airway obstruction. An overview of the study design is shown in Figure E1.

### Longitudinal Observational Study

#### Population

The NHLBI Pooled Cohorts Study harmonized spirometry data from nine U.S. population-based cohorts.^12^ Of these, four cohorts—the Atherosclerosis Risk in Communities (ARIC) Study, Coronary Artery Risk Development in Young Adults (CARDIA) Study, Cardiovascular Health Study (CHS), and the Multi-Ethnic Study of Atherosclerosis (MESA)— measured omega-3 fatty acids in plasma phospholipids in some or all participants. We included all NHLBI Pooled Cohorts Study participants identifying as Black, white, Asian, or Hispanic with plasma phospholipid omega-3 fatty acids and lung function measurements.

#### Omega-3 Fatty Acid Measurements

Details on omega-3 fatty acid measurements in each cohort are presented in Table E1. Concentrations of ALA, EPA, DPA and DHA were expressed as a percentage of total plasma phospholipid fatty acids.

#### Lung Function Measurements

FEV_1_ and FVC were measured by spirometry. Details on cohort-specific spirometry are shown in Table E1. Harmonization of spirometry data across cohorts was previously described.^12^ Airway obstruction was defined as FEV_1_/FVC < 70%, following the criteria for persistent airflow limitation described in the Global Initiative for Chronic Obstructive Lung Disease 2022 report.^13^

#### Statistical Analysis

Baseline characteristics were calculated for the overall sample and by cohort. Differences in omega-3 fatty acid concentrations between participants who did or did not develop airway obstruction during follow-up were evaluated with Student’s t-tests. Correlations between fatty acid concentrations were evaluated with Pearson’s correlations.

Associations of omega-3 fatty acids with FEV_1_ and FVC decline were evaluated with linear mixed effects modeling. Associations of omega-3 fatty acids with incident airway obstruction were evaluated with Cox proportional hazards modeling. Details on statistical models, including covariate adjustments, are provided in the online data supplement.

Effect modification by sex, race/ethnicity, and smoking was investigated with three-way interaction terms. The Benjamini and Hochberg false discovery rate correction for multiple testing was applied to interaction analyses separately for each lung function phenotype; both unadjusted and false discovery rate adjusted p-values are presented. Stratified estimates were derived from interaction terms and covariances.

Analyses were performed in SAS version 9.4.

#### MR Study

Two-sample MR was performed to estimate associations of genetically predicted EPA, DPA, DHA, and total omega-3 fatty acids with FEV_1_, FVC, and spirometry-defined airway obstruction.

#### Omega-3 Fatty Acid Genetic Instruments

Genetic instruments were based on single nucleotide polymorphisms (SNPs) in distinct genetic loci identified in published omega-3 fatty acid GWAS.^14-17^ The F-statistic, a measure of instrument strength and validity, was calculated for all instruments.^18^

#### SNP–Omega-3 Fatty Acid Associations

Primary analyses for each fatty acid used SNP—fatty acid associations from the GWAS explaining, to our knowledge, the greatest proportion of variation in that fatty acid.^14,15^ Analyses were replicated using SNP—fatty acid associations from additional, independent GWAS.^16,17^ Summary data for all SNP—fatty acid associations are shown in Tables E2–E5.

#### SNP–Lung Function Associations

Publicly available data from lung function GWAS in European ancestry UK Biobank participants were used for all analyses.^19,20^ Data were extracted through the MR-Base platform (study IDs ukb-b-11141 for FEV_1_, ukb-b-14713 for FVC, and ieu-b-106 for airway obstruction). Further details on the genetic instruments and the omega-3 fatty acid and lung function GWAS are presented in the online data supplement.

#### Statistical Analysis

MR analyses used the inverse variance weighted (IVW) method, which assumes pleiotropy is either nonexistent or balanced, and the MR-Egger, weighted-median, and weighted mode methods, which can provide valid estimates when the assumption of zero or balanced pleiotropy is violated.^21-24^ Analyses were performed in R Studio version 1.2.1335 using the “TwoSampleMR” R package (https://github.com/MRCIEU/TwoSampleMR).^25^

## RESULTS

### Longitudinal Analysis

#### Participant Characteristics

Analyses of lung function decline included 15,063 participants from four cohorts (Table 1). The mean age at first spirometry visit was 56 years. The total sample was comprised of 55% female and 69% white, 20% Black, 4% Asian, and 7% Hispanic participants. Forty-eight percent were never smokers, 36% were former smokers, and 16% were current smokers. Mean concentrations for ALA, EPA, DPA, and DHA were 0.2%, 0.7%, 0.9%, and 3.3%, respectively, of total plasma phospholipid fatty acids. Participants had a mean of 2.6 spirometry measurements; 82% had at least two measurements and 49% had at least three measurements. The median (interquartile range) follow-up time among participants with at least two spirometry measurements was 7(16) years. Twenty-three percent of participants had spirometry-defined airway obstruction at baseline. Characteristics of participants without airway obstruction at baseline are shown in Table E6.

**Table 1.** Characteristics of NHLBI Pooled Cohorts Study participants at first spirometry measurement.

|  | **Overall** | **ARIC** | **CARDIA** | **CHS** | **MESA** |
| --- | --- | --- | --- | --- | --- |
| **Participants, n (%)** | 15063 (100) | 3545 (24) | 3303 (22) | 3727 (25) | 4488 (30) |
| **Age, years** | 56.3 (18.8) | 54.7 (5.8) | 25.6 (3.9) | 72.5 (5.3) | 66.8 (9.9) |
| **Female sex, n (%)** | 8224 (55) | 1836 (52) | 1867 (57) | 2196 (59) | 2325 (52) |
| **Race/ethnicity, N (%)** |  |  |  |  |  |
| *white* | 10434 (69) | 3545 (100) | 1792 (54) | 3341 (90) | 1756 (39) |
| *Black* | 3008 (20) | -- | 1511 (46) | 383 (10) | 1114 (25) |
| *Asian* | 641 (4) | -- | -- | 3 (0) | 638 (14) |
| *Hispanic* | 980 (7) | -- | -- |  | 980 (22) |
| **Height, cm** | 167.4 (19.9) | 169.3 (9.5) | 170.3 (9.5) | 164.8 (9.3) | 165.8 (10) |
| **Weight, kg<sup>a</sup>** | 75.1 (16.5) | 77.9 (16.4) | 71.3 (16.1) | 72.3 (14.2) | 78 (17.6) |
| **Smoking status, n (%)** |  |  |  |  |  |
| *Never* | 7169 (48) | 1303 (37) | 2004 (61) | 1785 (48) | 2077 (46) |
| *Former* | 5440 (36) | 1422 (40) | 446 (14) | 1566 (42) | 2006 (45) |
| *Current* | 2454 (16) | 820 (23) | 853 (26) | 376 (10) | 405 (9) |
| **Smoking pack-years, median (IQR)<sup>b</sup>** | 15.3 (30.2) | 24.8 (27.0) | 2.3 (5.2) | 27.0 (36.4) | 12.2 (26.5) |
| **Smoking cigarettes per day, median (IQR)<sup>c</sup>** | 15 (10) | 20 (10) | 10 (14) | 19.5 (10) | 10 (15) |
| **FEV<sub>1</sub>, mL** | 2712 (930) | 3051 (780) | 3549 (796) | 2089 (656) | 2345 (732) |
| **FVC, mL<sup>d</sup>** | 3584 (1118) | 4113 (983) | 4303 (1032) | 2964 (867) | 3147 (955) |
| **FEV<sub>1</sub>/FVC < 0.7, n (%)<sup>b</sup>** | 3405 (23) | 787 (22) | 119 (4) | 1461 (39) | 1045 (23) |
| **Follow-up time, yr, median (IQR)<sup>e</sup>** | 4 (16.0) | 3.0 (20.1) | 20 (1.0) | 6.8 (3.0) | 11.0 (6.0) |
| **PFT measurements, n** | 2.6 (1.2) | 2.2 (0.6) | 4.3 (0.9) | 2.3 (0.7) | 2 (0.8) |
| **ALA, %<sup>f</sup>** | 0.2 (0.1) | 0.1 (0.1) | 0.2 (0.1) | 0.2 (0.1) | 0.2 (0.1) |
| **EPA, %<sup>f</sup>** | 0.7 (0.6) | 0.6 (0.3) | 0.8 (0.5) | 0.6 (0.4) | 0.9 (0.8) |
| **DPA, %<sup>f</sup>** | 0.9 (0.2) | 0.9 (0.2) | 0.9 (0.2) | 0.8 (0.2) | 0.9 (0.2) |
| **DHA, %<sup>f</sup>** | 3.3 (1.2) | 2.8 (0.9) | 3.2 (1.1) | 3.0 (1.0) | 3.9 (1.5) |

All values reported as mean (SD) unless otherwise noted.
<sup>a</sup> Six participants were missing baseline weight.
<sup>b</sup> Smoking pack-years calculated among former and current smokers (N = 7,894).
<sup>c</sup> Smoking cigarettes per day calculated among current smokers (N = 2,454).
<sup>d</sup> N = 15,022 for FVC and FEV<sub>1</sub>/FVC due to failure to meet spirometry reproducibility criteria.
<sup>e</sup> Follow-up time calculated among participants with at least two spirometry measurements (N = 12,375).
<sup>f</sup> Percent of total fatty acids in plasma phospholipids.

#### Correlations of omega-3 fatty acids

Correlations of ALA with EPA, DPA, and DHA were evaluated in the full sample and by sex, race/ethnicity, and smoking status (Table E7). In the full sample, ALA was positively correlated with EPA, DPA, and DHA, with decreasing magnitudes (Pearson’s correlation coefficients [r] = 0.18, 0.10, and 0.03 for EPA, DPA, and DHA respectively, p < 0.0001 for all). Correlation patterns were similar in male participants, white participants, and never and former smokers. In contrast, ALA and DHA were not correlated in female participants (r = 0.01, p = 0.255) and were negatively correlated in Black participants (r = -0.04, p = 0.034), Hispanic participants (r = -0.09, p = 0.004), and current smokers (r = -0.04, p = 0.067).

#### Lung Function Decline

The mean rate of lung function decline was 36.8 mL per year for FEV_1_ and 35.8 mL per year for FVC. In covariate-adjusted models, higher concentrations of ALA, EPA, and DHA were associated with attenuated decline for both FEV_1_ and FVC (Table 2). One SD higher ALA, EPA, and DHA was associated with an attenuation of 0.77, 0.62, and 1.77 mL/year for FEV_1_ and 1.30, 0.84, and 2.43 mL/year for FVC (p < 0.005 for all). In extended models that accounted for the effects of sex, smoking, and race/ethnicity on lung function decline, the associations of ALA with FEV_1_ decline and of EPA with both FEV_1_ and FVC decline were no longer statistically significant (p > 0.05), while the association of DPA with FEV_1_ decline was strengthened (p = 0.03). The associations of DHA with FEV_1_ and FVC decline remained statistically significant (p < 0.05) in all models.

**Table 2.**
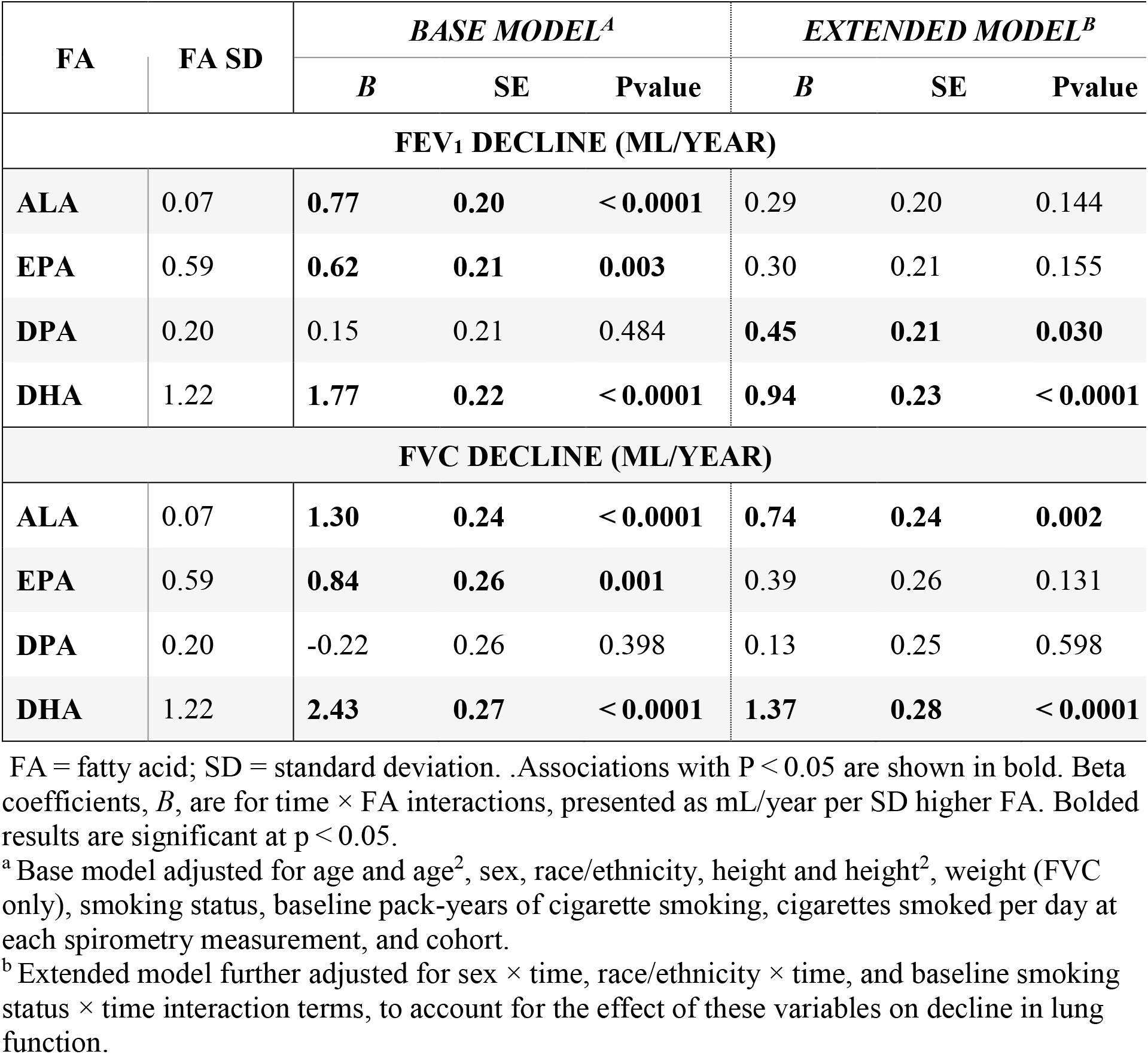
Associations of omega-3 fatty acids with FEV_1_ and FVC decline.

Three-way interaction terms revealed nominally statistically significant modification by sex, smoking, and/or race/ethnicity (interaction p < 0.05) of some omega-3 fatty acid—lung function decline associations (Table E8). All interaction terms remained significant after correction for multiple testing (adjusted p < 0.05), except for the “sex by ALA by elapsed time” and the “race by EPA by elapsed time” interaction terms for FEV_1_ (adjusted p = 0.053 and 0.059, respectively).

Where there was evidence for differences by biological sex, associations were observed primarily in male participants. For example, one SD higher ALA was associated with attenuated FEV_1_ decline of 1.01 (95% confidence interval [CI] = 0.32–1.71) mL/year in males; there was little to no association in females (beta = 0.08, 95% CI = -0.38–0.54) (Figure 1A). Similar sex-specific patterns were observed for associations of EPA and DPA with FEV_1_ decline (Figure 1A and 1B) and ALA with FVC decline (Figure E2A).

**Figure 1.**
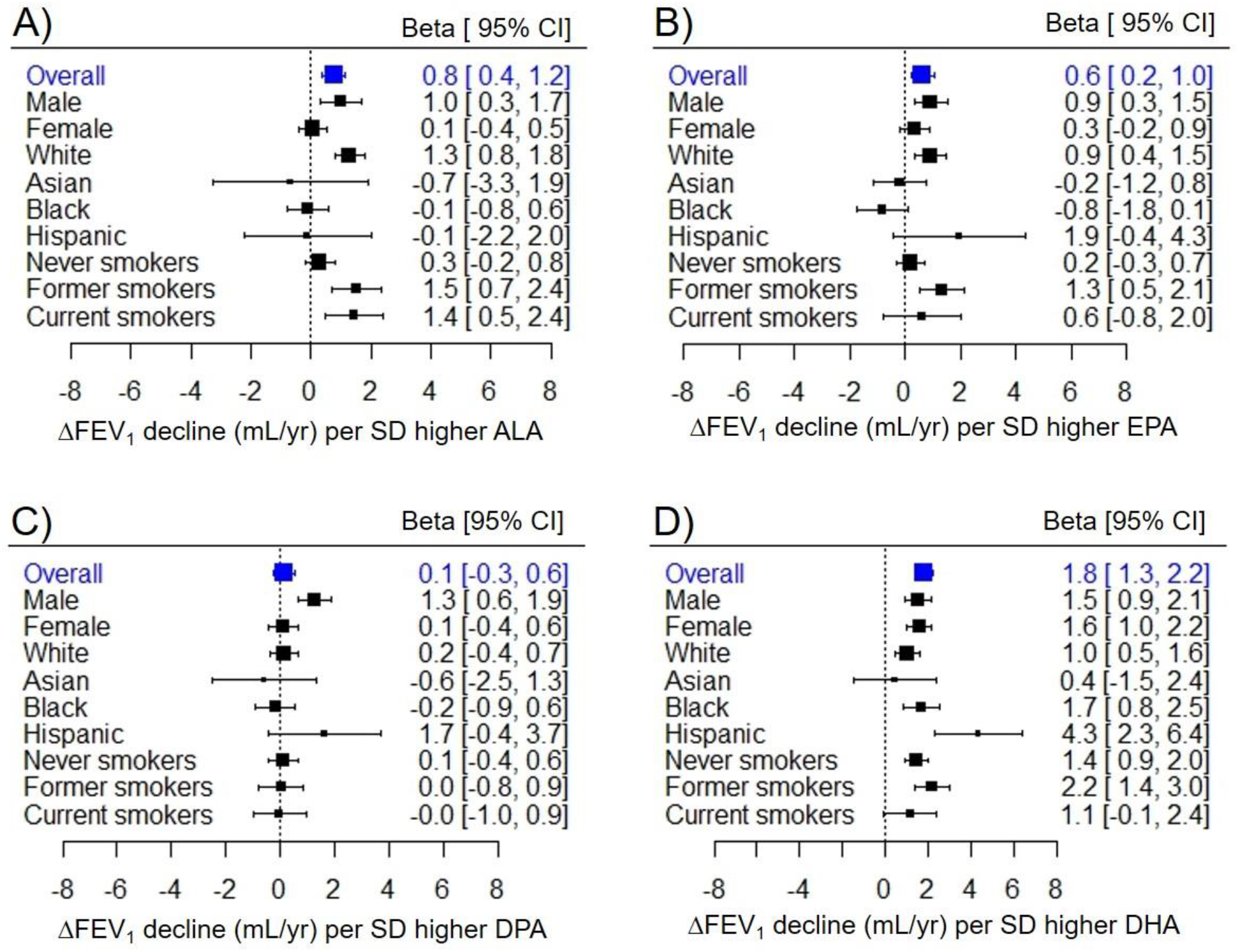
Stratification of omega-3 fatty acid–FEV_1_ decline associations by sex, race/ethnicity, and baseline smoking status. Sex, race/ethnicity, and baseline smoking status differences in omega-3 fatty acid–FEV_1_ decline associations for A) ALA, B) EPA, C) DPA, and D) DHA. Estimates (Betas) and 95% confidence intervals (CIs) for each group were derived from three-way interaction terms and covariances. All estimates are presented per standard deviation (SD) higher fatty acid.

Where there was evidence for differences by smoking, associations were observed primarily or strongest in former smokers. For example, one SD higher EPA was associated with attenuated FEV_1_ decline of 1.3 mL/year (95% CI = 0.5–2.1) in former smokers, 0.6 mL/year (95% CI = -0.8–2.0) in current smokers, and 0.2 mL/year (95% CI = -0.3–0.7) in never smokers (Figure 1B). There were similar patterns for associations of ALA and DHA with FEV_1_ decline (Figure 1A and 1D) and for ALA, EPA and DHA with FVC decline (Figure E2A, E2B and E2D).

Effect modification by race/ethnicity varied depending on the omega-3 fatty acid. The positive associations of ALA and EPA with lung function decline were observed primarily in white participants (Figure 1A and 1B). In contrast, the positive associations of the DHA with lung function decline were largest in Black and Hispanic participants, still positive and statistically significant in white participants, and non-significant in Asian participants (Figure 1D). Race/ethnicity specific results for FVC decline followed a similar pattern (Figure E2).

#### Incident Airway Obstruction

There were 1,455 (12.5%) cases of incident airway obstruction during a median (interquartile range) follow-up period of 5(6) years. Characteristics of cases and participants who did not develop airway obstruction during follow-up are shown in Table E6. Concentrations of ALA, EPA and DPA were similar between cases and those who did not develop airway obstruction, while DHA concentration was slightly lower in cases (3.23% vs 3.29% of total fatty acids, P_diff_ = 0.05).

In covariate-adjusted models, higher ALA was associated with higher rates of airway obstruction (hazard ratio = 1.08, 95% CI = 1.03–1.13), while higher DHA was associated with lower rates of airway obstruction (hazard ratio = 0.91, 95% CI = 0.87–0.97) (Figure 2). When all omega-3 fatty acids were included in the same model, the association of ALA with rates of incident airway obstruction was attenuated (hazard ratio = 1.04, 95% CI = 1.00–1.08), while the association of DHA with rates of incident airway obstruction was unchanged (hazard ratio = 0.91, 95% CI = 0.85–0.98).

**Figure 2.**
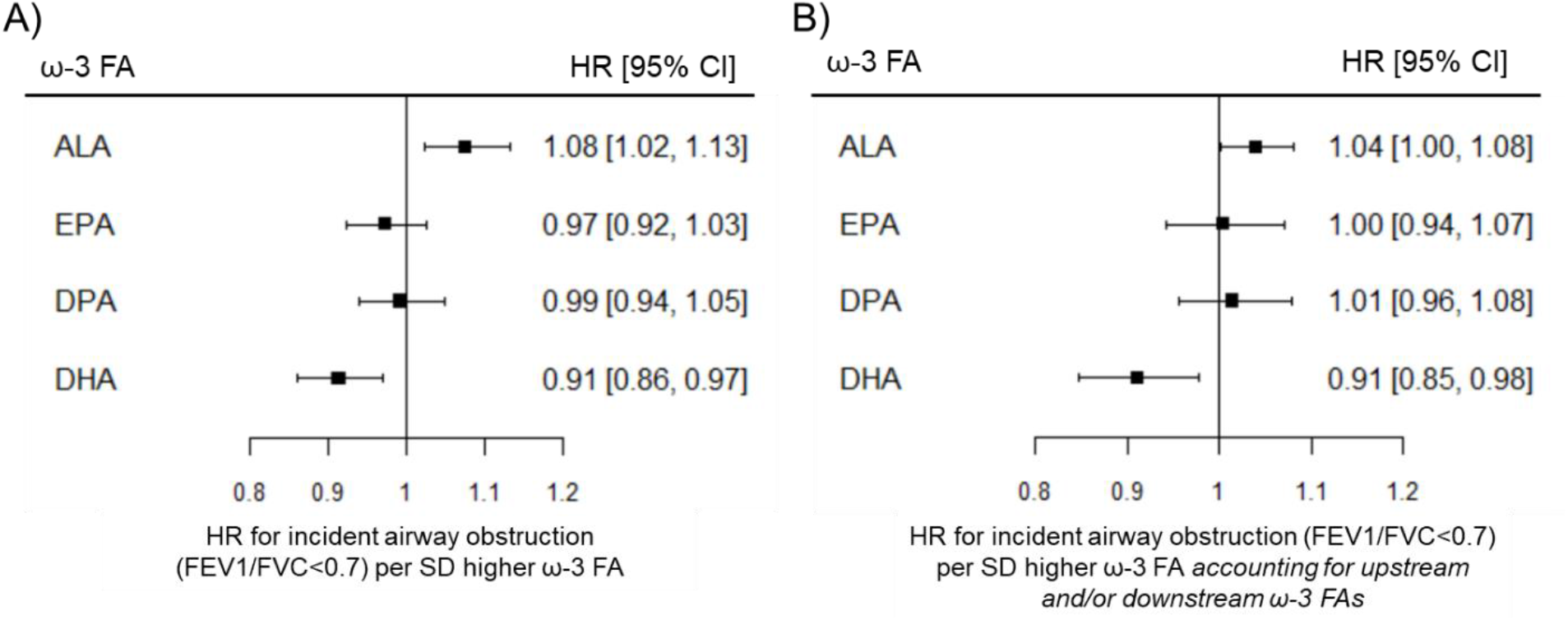
Associations of omega-3 fatty acids with incident airway obstruction in participants without airway obstruction at baseline. Estimates from Cox proportional hazards models. HR = hazard ratio. CI = confidence interval. SD = standard deviation. A) Associations of omega-3 fatty acids with incident airway obstruction adjusted baseline age, sex, race/ethnicity, height, smoking status, pack-years of cigarette smoking, and cohort. B) Associations of omega-3 fatty acids with incident airway obstruction further adjusted for other upstream and/or downstream omega-3 fatty acids.

There was little evidence for associations of EPA or DPA with incident airway obstruction. There was no statistically significant evidence for effect modification by sex, race/ethnicity, or smoking for any omega-3 fatty acid (Table E7).

#### Two-Sample MR Analysis

##### Genetic Instruments for Omega-3 Fatty Acids

Genetic instruments for EPA, DPA, DHA, and total omega-3 fatty acids explained 2.1%, 8.5%, 6.5%, and 7.4% of the variance, respectively. In the primary analyses, F-statistics for the omega-3 fatty acid genetic instruments ranged from 97 for EPA to 275 for DPA, exceeding the MR standard for instrument strength of F-statistic > 10 (Figure 3). F-statistics were generally lower in the replication analyses, ranging from 6 for DHA to 44 for total omega-3 fatty acids (Table E9).

**Figure 3.**
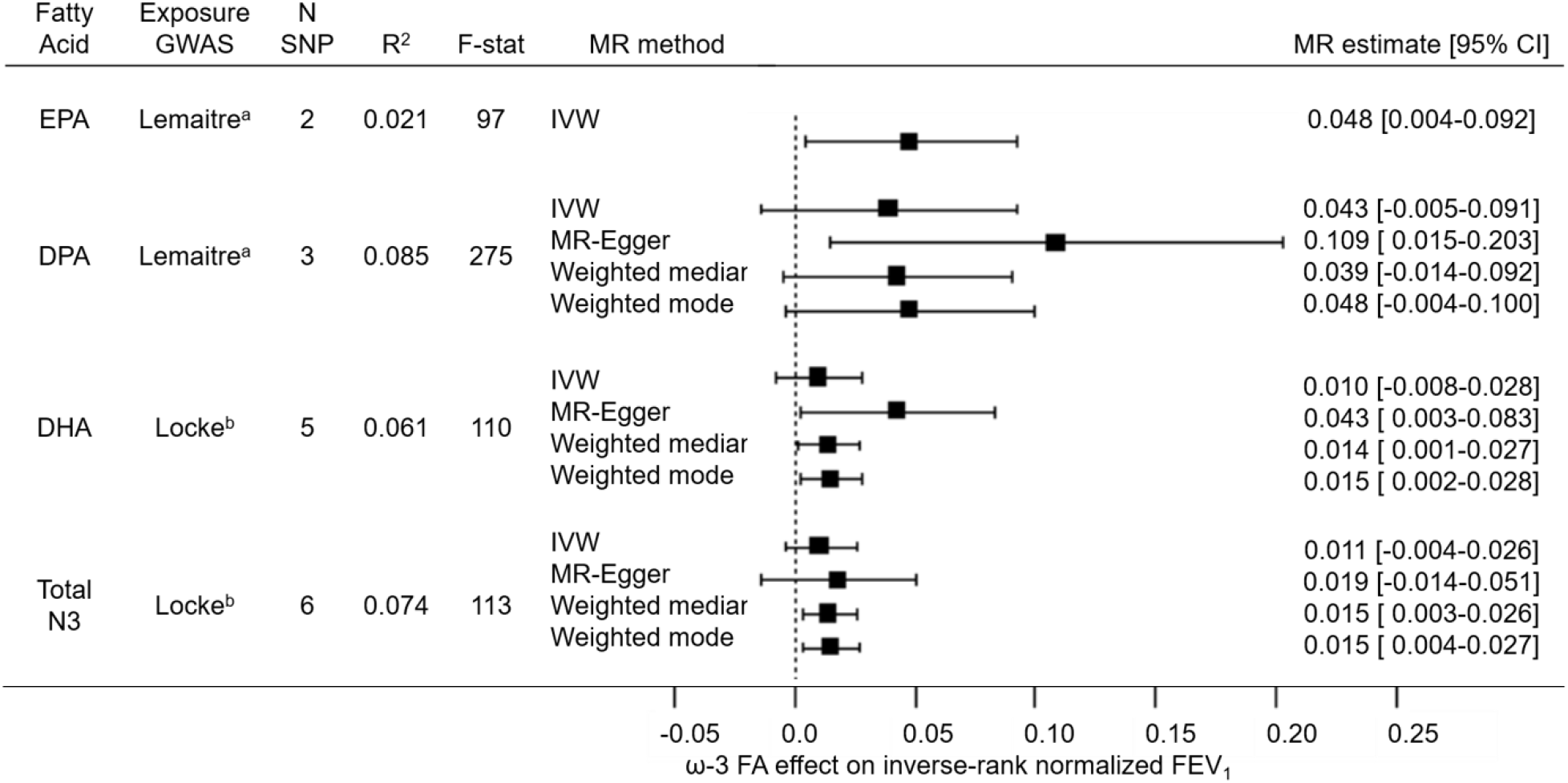
Forest plot of two-sample MR estimates from primary analysis of EPA, DPA, and DHA and total omega-3 fatty acids on FEV_1_. Forest plot showing MR estimates and 95% confidence intervals (CIs) for associations of genetically predicted omega-3 fatty acids with FEV_1_ using different MR methods. N SNP: number of SNPs contributing to the omega-3 fatty acid genetic instrument. Exposure GWAS: GWAS that SNP–omega-3 fatty acid association data were extracted from. R^2^: proportion of omega-3 fatty acid variance explained by the genetic instrument. F-stat: F statistic, a measure of genetic instrument strength where the MR standard for validity is F > 10. IVW: Inverse variance weighted MR. ^a^ Lemaitre study: GWAS fatty acid units in % of total fatty acid composition. ^b^ Locke study: GWAS fatty acid units in standardized fatty acid concentration (mmol/L).

##### MR of Omega-3 Fatty Acids and Lung Function

Associations of genetically predicted omega-3 fatty acids with FEV_1_ were consistently positive across different MR methods (Figure 3). Results were statistically significant (p < 0.05) for the following: IVW estimate for EPA; the MR-Egger estimate for DPA; the MR-Egger, weighted median, and weighted mode methods for DHA; and the weighted median and weighted mode methods for total omega-3 fatty acids. Results for FVC were also consistently positive, with statistically significant results (p <0.05) for the MR-Egger estimate for DPA (Figure E3). There was no evidence for an association of genetically predicted omega-3 fatty acids with the odds of airway obstruction (Figure E4).

Results were similar in replication analyses (Table E9). MR estimates for FEV_1_ and FVC were consistently positive across different methods, with statistically significant results (p <0.05) for some methods. Again, there was no statistically significant evidence for associations of genetically predicted omega-3 fatty acids with the odds of airway obstruction.

## DISCUSSION

Our longitudinal pooled individual participant data analysis in a large, multiracial U.S. population-based sample found that higher levels of omega-3 fatty acids in circulation were associated with an attenuated rate of lung function decline, with the strongest associations seen for DHA. Higher DHA was also associated with decreased risk of incident airway obstruction. Results from the MR study, although not significant across all MR methods, were consistent with a positive association of genetically predicted omega-3 fatty acids with lung function. Together, the findings from the longitudinal and MR studies support a protective effect of higher circulating omega-3 fatty acids—most significantly DHA—on lung health.

Circulating omega-3 fatty acids correlate with lung function parameters cross-sectionally,^5, 6, 8, 9^ and longitudinal studies of dietary intake have shown protective associations of higher omega-3 fatty acid intake with lung function decline and the odds of COPD.^26, 27^ To our knowledge, this is the first study to longitudinally evaluate the association of omega-3 fatty acid biomarkers with lung function decline in a generally healthy adult population, and the first study to apply MR methodology to the omega-3 fatty acid–lung function relationship. Triangulation of our longitudinal and MR findings helps to address limitations of reverse causality and confounding in previous studies, and contributes to the evidence base for a beneficial effect of omega-3 fatty acids on long-term lung outcomes. Beneficial effects of omega-3 FAs in the lungs have also been shown experimentally.^28-32^ In vitro, the omega-3 fatty acid-derived small lipid molecule, resolvin E1, reduced the production of proinflammatory cytokines and stress-related proteins in human alveolar epithelial cells.^28^ In animal studies, dietary or pharmacological supplementation with omega-3 fatty acids protected against experimentally-induced airway inflammation and resulting lung pathology, and accelerated lung tissue recovery following lung injury.^29-32^ In clinical studies of acute lung injury, omega-3 fatty acid supplementation decreased inflammation in the lungs and accelerated respiratory function recovery, as recently reviewed.^33^ Altogether, the anti-inflammatory effects of omega-3 fatty acids in the lungs are well established, providing strong biologic plausibility that support our findings.

Previous studies of omega-3 fatty acids and lung outcomes have not consistently investigated effect modification by important biological and lifestyle factors. In our longitudinal study, we investigated whether associations of omega-3 fatty acids with lung function decline and incident airway obstruction differed by sex, smoking, and race/ethnicity, and found both qualitative and quantitative differences. We found that the positive associations of ALA, EPA, and DPA with lung function decline was observed primarily in males, white participants, and/or those with a history of smoking. In contrast, the positive associations of DHA with lung function decline was observed in males and females; in Black, white and Hispanic participants; and in all smoking groups, with the largest magnitude associations in Black and Hispanic participants and former smokers. Correlations between fatty acid levels also differed by sex, race/ethnicity, and smoking. ALA and DHA were positively correlated in males, white participants, and former smokers, but were uncorrelated or negatively correlated in females, Black and Hispanic participants, and current smokers. These patterns are consistent with reported differences in the efficiency of omega-3 fatty acid metabolism by sex, race/ethnicity, and smoking.^34-41^ Unfortunately, we were unable to test these differences in the MR analyses because omega-3 fatty acid GWAS stratified by sex or smoking are unavailable and the limited sample sizes of lung function GWAS of non-European populations are underpowered for MR analyses.

In our longitudinal study, the protective associations of omega-3 fatty acids with incident airway obstruction were limited to DHA, with little evidence for differences by sex, smoking status, or race/ethnicity. In our MR study, we found no evidence for associations of omega-3 fatty acids with odds of airway obstruction. The weaker associations and lack of interaction effects for the omega-3 fatty acid–airway obstruction associations could be explained by a loss of power due to assessment of a binary (vs. continuous) outcome and, for the longitudinal study, smaller sample size (limited to participants without airway obstruction at baseline). Larger sample sizes, more precise COPD phenotyping, and increased statistical power may be necessary to detect long-term effects of omega-3 fatty acids on obstructive airway disease.

This study has several strengths and novel aspects. Our longitudinal study pooled individual participant data from multiple cohorts, leading to a large, diverse sample with repeated lung function measurements (up to 25 years of follow-up in some participants). These data also enabled investigation of effect modification by important biological and lifestyle factors. For the exposure, we used omega-3 fatty acids biomarkers, which reflect dietary intake while minimizing reporting bias and measurement error in dietary intake assessed from diet recalls or food frequency questionnaires. Similar or identical laboratory methods were used to measure omega-3 fatty acids across the cohorts, increasing confidence that the data were suitable for pooling. Our MR study used publicly available GWAS summary statistics, which allowed us to maximize sample size, instrument strength, and statistical power. Our MR findings were consistent in direction across multiple MR methods designed to address potential violation in the assumptions of MR and support our longitudinal findings.

Our analyses also have limitations. For our longitudinal study, plasma phospholipids, which represent short-term fatty acid status, were measured only once in each cohort. Multiple spirometry measurements were taken in all cohorts, but the number of spirometry measurements and intervals between spirometry measurements varied. Repeated omega-3 fatty acids generally correlate over time, but one-time measurements may not reflect fluctuations in omega-3 fatty acids bioavailability due to dietary or lifestyle changes, especially in participants with longer total follow-up times. Our findings may have been strengthened if repeated measurements of omega-3 fatty acids and additional spirometry measurements were taken at consistent intervals over longer follow-up periods across the cohorts. This may be especially true for the analysis of incident airway obstruction, since censoring and the identification of cases in each cohort was limited to the cohort clinic visits where spirometry was performed. Additionally, omega-3 fatty acids were reported as percentages of total plasma phospholipids rather than absolute concentrations. Fatty acids proportions and absolute concentrations are generally well correlated, suggesting the use of relative versus absolute concentrations is unlikely to influence interpretation of our results.^42, 43^ Finally, our sample had limited numbers of Asian and Hispanic participants; although we found significant positive associations of DHA with lung function decline in Hispanic participants, our analyses may have been underpowered to detect other associations in these groups. For our MR study, the genetic instruments for the different omega-3 fatty acids included SNPs in genetic loci associated with multiple fatty acids, limiting the specificity of the results. Our analysis relied on publicly available GWAS summary statistics; stratified GWAS of omega-3 FAs were not available and that limited our ability to confirm the sex- and smoking-specific effects seen in our longitudinal cohort study. Our MR study was also limited to European ancestry; larger GWAS of omega-3 FAs and lung function parameters, specifically in non-European populations, are needed to confirm our longitudinal findings.

Overall, this study represents the first longitudinal study of omega-3 fatty acid biomarkers and lung function decline and the first MR study of genetically predicted omega-3 fatty acids and lung function parameters. Our findings are consistent in direction across the longitudinal and MR studies and provide evidence supporting a protective effect of omega-3 fatty acids—specifically DHA—on lung health. Further research into biological and lifestyle factors influencing omega-3 fatty acid metabolism and utilization in the lungs is warranted.

## Supporting information

STROBE Checklists

## Data Availability

For the longitudinal cohort study, pre-existing data access policies for each of the parent cohort studies specify that research data requests can be submitted to each steering committee; these will be promptly reviewed for confidentiality or intellectual property restrictions and will not unreasonably be refused. Individual-level patient data may be further restricted by consent, confidentiality or privacy laws/considerations.
For the Mendelian Randomization study, all analyses used publically available summary data, and all results produced are contained in the manuscript.

## ACKNOWLEDGMENTS

The authors thank the staff and participants of the Atherosclerosis Risk in Communities Study, the Coronary Artery Risk Development in Young Adults Study, the Cardiovascular Health Study, and the Multi-Ethnic Study of Atherosclerosis for their important contributions.

