## Supplementary material for "Investigating associations of omega-3 fatty acids, lung function decline, and airway obstruction": Online Data Supplement

**Supplemental Methods**

*Longitudinal study statistical models*

Associations of omega-3 fatty acid concentrations with lung function decline were evaluated with linear mixed effects modeling of repeated lung function measurements regressed on omega-3 fatty acid levels, elapsed time since first lung function measurement, and the omega-3 fatty acid × elapsed time multiplicative interaction term. Models were run separately for the four omega-3 fatty acids (ALA, EPA, DPA and DHA) on decline in two spirometry measures ($FEV_1$ and FVC), and adjusted for age and $age^2$, sex, self-reported race/ethnicity, height and $height^2$, weight (FVC only), smoking status (current vs. former vs. never) at each spirometry measurement, baseline pack-years of cigarette smoking, cigarettes smoked per day at each spirometry measurement, and cohort. All covariates were specified as fixed effects. To allow variation across participants in baseline lung function and lung function decline over follow-up, the intercept and elapsed time parameters were specified as random effects. Extended models included multiplicative interaction terms of sex, self-reported race/ethnicity, and smoking status with elapsed time to account for the associations of these covariates on lung function decline. Results are shown as mL/year per SD increase in omega-3 fatty acid % of total plasma phospholipid fatty acids.

Associations of omega-3 fatty acids with incident airway obstruction, defined as $FEV_1/FVC < 70\%$, were evaluated with Cox proportional hazards modeling. Analyses were limited to participants with $FEV_1/FVC \geq 70\%$ at first spirometry measurement. Elapsed time since first spirometry measurement was treated as time to event. Participants with $FEV_1/FVC \geq 70\%$ at their last spirometry measurement were right-censored. Analyses were adjusted for baseline age, sex, race/ethnicity, height, baseline smoking status, baseline pack-years of cigarette smoking, and cohort.

*Mendelian Randomization study genetic instruments*

Genetic instruments for omega-3 fatty acids were based on distinct genetic loci identified in published omega-3 fatty acid GWAS. Distinct genetic loci were defined with the PLINK clumping method with a linkage-disequilibrium (LD) threshold of $r^2 = 0.01$. For each fatty acid, the single nucleotide polymorphisms (SNPs) comprising the genetic instrument were defined by the results of the GWAS that explained, to our knowledge, the greatest proportion of variation in that fatty acid. Genetic instruments for EPA and DPA were defined by results from a meta-analysis GWAS of plasma phospholipid omega-3 fatty acids in the Cohorts for Heart and Aging Research in Genomic Epidemiology (CHARGE) consortium and explained 2.1% and 8.5% of the variance, respectively.[1] Genetic instruments for DHA and total omega-3 fatty acids were defined by a GWAS of quantitative traits in the Metabolic Syndrome in Men (METSIM) and National FINRISK studies and explained 6.5% and 7.4% of the variance, respectively.[2]

*Mendelian Randomization study SNP-omega-3 fatty acid associations*

Summary data for SNP–fatty acid associations were obtained for each SNP contributing to the EPA, DPA, DHA, and total omega-3 fatty acid genetic instruments (Tables E2–E5). For EPA and DPA, the primary analysis used results from the CHARGE meta-analysis GWAS (N = 8,866), and the replication analysis used results from a meta-analysis GWAS of circulating metabolites in the TwinsUK and Cooperative Health Research in the Region Augsburg (KORA) studies (N = 7,816 for EPA and 7,821 for DPA).[3] For DHA and total omega-3 fatty acids, the primary analysis used results from the METSIM/FINRISK GWAS (N = 8,751), and the replication analysis used results from the CHARGE GWAS (DHA only, N = 8,866), the TwinsUK/KORA meta-analysis GWAS (DHA only, N = 7,818), and a meta-analysis GWAS of circulating metabolites in 14 European cohorts (N = 13,499 for DHA and N = 13,544 for total omega-3 fatty acids).[4] The details of the four omega-3 fatty acid GWAS have been previously described.[1-4]

*Mendelian Randomization SNP-lung function associations*

Data from the PHEASANT GWAS pipeline were used for $FEV_1$ and FVC (respective study IDs ukb-b-11141 and ukb-b-14713; N = 345,665 for both).[5] Data from the Medical Research Council Integrative Epidemiology Unit UK Biobank GWAS pipeline were used for spirometry-defined airway obstruction (study ID ieu-b-106; N = 353,315, including 55,907 cases and 297,408 controls).[6]

**Table E1. Details of omega-3 fatty acid and spirometry measurements in each cohort**

| Cohort | Cohort description | Omega-3 fatty acid measurements | Spirometry |
|---|---|---|---|
| Atherosclerosis Risk in Communities (ARIC) | The ARIC study is a population-based longitudinal cohort of about 15,800 primarily Black and white middle-aged men and women, recruited from four communities in the U.S. between 1987 and 1989. For this study, only participants from the suburban Minneapolis, MN, area were included, given that omega-3 fatty acid biomarker measures were conducted only in this ARIC subset.[7] | Fasting blood was collected at the first clinic visit (1987–1989) and plasma was stored at -70℃ until analysis. Lipid fractions were extracted with chloroform/methanol and separated using thin-layer chromatography (TLC). Methyl esters of fatty acids in plasma phospholipids were measured by gas chromatography (Model 5890, Hewlett-Packard, Avondale, PA). A total of 28 fatty acids were identified and the relative amount of each fatty acid (% of total fatty acids) was quantified.[7] | Spirometry was performed at clinic visits 1 (1987–1989), 2 (1990–1992), and 5 (2011–2013). Collins Survey II water-sealed spirometers (Warren E. Collins Inc., Braintree, MA) were used for visits 1 and 2, and SensorMedics model 1022 dry rolling seal spirometers (OMI, Houston, TX) were used for visit 5.[8] |
| Coronary Artery Risk Development in Young Adults (CARDIA) | The CARDIA study is a population-based longitudinal cohort of about 5,115 Black and white young adults aged 18 to 30, recruited from four U.S. metropolitan areas: Birmingham, AL, Chicago, IL, Minneapolis, MN, and Oakland, CA. CARDIA began in 1985–1986.[9] | Fasting blood was collected at exam year 20 (2005–2006) and plasma was frozen at -70℃. Lipids fractions were extracted with chloroform/methanol and separated using TLC. Fatty acid methyl esters were generated from transesterification from the phospholipid fraction and were measured by GC with a flame ionization detector. Twenty-eight fatty acids were identified, and the relative amount of each fatty acid (% of total fatty acids) was quantified.[1] | Spirometry was performed at baseline (1985–1986) and exam years 2 (1987–1988), 5 (1990–1991), 10 (1995–1996) and 20 (2005–2006). Collins Survey II water-sealed spirometers were used at baseline and exam years 2, 5 and 10, and SensorMedics model 1022 dry rolling seal spirometers were used at exam year 20.[8] |
| Cardiovascular Health Study (CHS) | CHS is a population-based longitudinal study of 520 men and women ≥ 65 years of age, recruited from four U.S. communities: Forsyth County, NC, Sacramento County, CA, Washington County, MD, and | Fasting blood samples were collected at examination 5 (1992–1993), and plasma was stored at -80℃. Plasma phospholipids were separated from other lipids fractions via 1-dimensional TLC. Phospholipid fatty acids were trans-methylated and measured by GC (5890, Agilent Technologies, Palo Alto, CA) | Spirometry was performed at cohort examination 2 (1989–1990), 6 (1993–1994), 9 (1996–1997) and 18 (2005–2006). Collins Survey II water-sealed spirometers were used at exams 2, 6, and 9, and EasyOne flow- |

| | | | |
|---|---|---|---|
| | Allegheny County (Pittsburgh), PA, in 1989. An additional 687 Black men and women were recruited starting in 1992. | with a SP-2560 fused-silica 100-m capillary column and a flame ionization detector. A total of 45 fatty acids were identified, and the relative amount of each fatty acid (% of total fatty acids) was quantified.[10] | sensing spirometers (ndd Medical Technologies, Inc., Andover, MA) were used for exam 18. For this study, the exam 9 data for FVC were excluded due to concerns regarding comparability between the FVC measures at exams 6 and 9, as recommended by CHS investigators.[11] |
| Multi-Ethnic Study of Atherosclerosis (MESA) | MESA is a population-based cohort consisting of 6,814 Black, white, Hispanic, and Chinese-American men and women 45 to 84 years of age, recruited from six sites in the U.S., including St. Paul, MN, Los Angeles, CA, northern Manhattan, NY, Forsyth County, NC, Chicago, IL, and Baltimore City and County, MD, from July 2000 to August 2002.[12,13] | Fasting blood samples were collected at study baseline (2000–2002) and plasma was stored at -70 °C. Plasma lipid fractions were extracted with chloroform/methanol and separated by TLC. The fatty acids in plasma phospholipids were trans-methylated and measured via GC (Model 5890, Hewlett-Packard, Avondale, PA) with a single capillary Varian CP7420 E15 100-m column with a flame ionization detector. The concentration was expressed as % of total fatty acids.[14] | Spirometry was performed at examination 3 or 4 (2004–2007), 5 (2010–2011), and 6 (2016–2017). SensorMedics model 1022 dry rolling seal spirometers were used for all exams.[8] |

**Table E2. EPA genetic instrument summary statistics**

| | | | Primary Analysis GWAS | | | | Replication Analysis GWAS | | | |
|---|---|---|---|---|---|---|---|---|---|---|
| | | | CHARGE (n = 8,866) Lemaitre et al. 2011 | | | | Twins UK/KORA (n = 7,816) Shin et al. 2014 | | | |
| **SNP** | **EA** | **NEA** | **EAF** | **BETA** | **SE** | **$R^2$** | **EAF** | **BETA** | **SE** | **$R^2$** |
| rs174538 | A | G | 0.280 | -0.083 | 0.005 | 0.018 | 0.303 | -0.034 | 0.004 | NA |
| rs3798713 | C | G | 0.430 | 0.035 | 0.005 | 0.004 | 0.423 | 0.004 | 0.004 | NA |

EA: effect allele. NEA: non-effect (other) allele. EAF: effect allele frequency. BETA: beta-coefficient for the effect allele on EPA, with fatty acid units defined by source GWAS (Lemaitre: % of total fatty acids; Shin: median-normalized and $\log_{10}$ transformed). $R^2$: proportion of variance EPA explained by each SNP, calculated using the equation $R^2_{snp} = 2\alpha^2$ MAF(1−MAF), where $\alpha$ = the standardized SNP-EPA association and MAF = minor allele frequency. For the Lemaitre et al. GWAS, betas were standardized by dividing the reported beta by the EPA standard deviation (0.4) prior to calculation of $R^2$. For the Shin et al. GWAS, $R^2$ could not be calculated due to incompatibility of the fatty acid units.

**Table E3. DPA genetic instrument summary statistics**

| | | | Primary Analysis GWAS | | | | Replication Analysis GWAS | | | |
|---|---|---|---|---|---|---|---|---|---|---|
| | | | CHARGE (n = 8,866) Lemaitre et al. 2011 | | | | Twins UK/KORA (n = 7,821) Shin et al. 2014 | | | |
| **SNP** | **EA** | **NEA** | **EAF** | **BETA** | **SE** | **$R^2$** | **EAF** | **BETA** | **SE** | **$R^2$** |
| rs174547 | C | T | 0.330 | -0.075 | 0.003 | 0.062 | 0.331 | -0.025 | 0.004 | NA |
| rs3734398 | C | T | 0.430 | 0.040 | 0.003 | 0.020 | 0.430 | 0.003 | 0.003 | NA |
| rs780094 | C | T | 0.590 | -0.017 | 0.003 | 0.003 | 0.603 | -0.008 | 0.003 | NA |

EA: effect allele. NEA: non-effect (other) allele. EAF: effect allele frequency. BETA: beta-coefficient for effect of the effect allele on DPA, with fatty acid units defined by source GWAS (Lemaitre: % of total fatty acids; Shin: median-normalized and $\log_{10}$ transformed). $R^2$: proportion of variance in DPA explained by each SNP, calculated using the equation $R^2_{snp} = 2\alpha^2$ MAF(1−MAF), where $\alpha$ = the standardized SNP-DPA association and MAF = minor allele frequency. For the Lemaitre et al. GWAS, betas were standardized by dividing the reported beta by the DPA standard deviation (0.2) prior to calculation of $R^2$. For the Shin et al. GWAS, $R^2$ could not be calculated due to incompatibility of the fatty acid units.

**Table E4. DHA genetic instrument summary statistics**

| | | | Primary Analysis GWAS | | | | Replication Analysis GWAS | | | | | | | | | | | |
|---|---|---|---|---|---|---|---|---|---|---|---|---|---|---|---|---|---|---|
| | | | METSIM/FINRISK (n = 8,751) Locke et al. 2019 | | | | 13 European Cohorts (n = 13,499) Kettunen et al. 2016 | | | | Twins UK/KORA (n = 7,818) Shin et al. 2014 | | | | CHARGE (n = 8,866) Lemaitre et al. 2011 | | | |
| **SNP** | **EA** | **NEA** | **EAF** | **BETA** | **SE** | **$R^2$** | **EAF** | **BETA** | **SE** | **$R^2$** | **EAF** | **BETA** | **SE** | **$R^2$** | **EAF** | **BETA** | **SE** | **$R^2$** |
| rs174547 | C | T | 0.43 | -0.293 | 0.016 | 0.042 | 0.40 | -0.127 | 0.012 | 0.008 | 0.33 | -0.009 | 0.003 | NA | 0.317 | -0.069 | 0.015 | 0.001 |
| rs2228603 | T | C | 0.06 | -0.213 | 0.033 | 0.005 | 0.07 | -0.130 | 0.025 | 0.002 | 0.08 | -0.015 | 0.006 | NA | 0.078 | -0.017 | 0.028 | 0.000 |
| rs187429064 | G | A | 0.06 | -0.202 | 0.034 | 0.005 | 0.04 | -0.204 | 0.038 | 0.003 | NA | | | | NA | | | |
| rs2229738 | T | C | 0.17 | -0.135 | 0.021 | 0.005 | 0.14 | -0.076 | 0.022 | 0.001 | NA | | | | NA | | | |
| rs75306442 | T | C | 0.19 | -0.108 | 0.020 | 0.004 | 0.19 | -0.051 | 0.017 | 0.001 | NA | | | | NA | | | |

EA: effect allele. NEA: non-effect (other) allele. EAF: effect allele frequency. BETA: beta-coefficient for effect of the effect allele on DHA, with fatty acid units defined by source GWAS (Locke and Kettunen: inverse-normal transformed; Shin: median-normalized and $\log_{10}$ transformed; Lemaitre: % of total fatty acids). $R^2$: proportion of variance in DHA explained by each SNP, calculated using the equation $R^2_{snp} = 2\alpha^2$ MAF(1−MAF), where $\alpha$ = the standardized SNP-DHA association and MAF = minor allele frequency. For the Lemaitre et al. GWAS, betas were standardized by dividing the reported beta by the DHA standard deviation (1.25) prior to calculation of $R^2$. For the Shin et al. GWAS, betas could not be converted and $R^2$ was unable to be calculated. rs187429064, rs2229738, and rs75306442 were not available in Shin et al. and Lemaitre et al. GWAS, and no suitable proxies (LD $r^2 > 0.8$) were identified.

**Table E5. Total omega-3 fatty acid genetic instrument summary statistics**

| | | | Primary Analysis GWAS | | | | Replication Analysis GWAS | | | |
|---|---|---|---|---|---|---|---|---|---|---|
| | | | METSIM/FINRISK (n = 8,751) Locke et al. 2019 | | | | 13 European Cohorts (n = 13,544) Kettunen et al. 2016 | | | |
| **SNP** | **EA** | **NEA** | **EAF** | **BETA** | **SE** | **$R^2$** | **EAF** | **BETA** | **SE** | **$R^2$** |
| rs10882229 | G | A | 0.376 | -0.083 | 0.016 | 0.003 | 0.383 | -0.009 | 0.013 | 0.000 |
| rs174546 | T | C | 0.426 | -0.323 | 0.016 | 0.051 | 0.403 | -0.154 | 0.012 | 0.011 |
| rs187429064 | G | A | 0.060 | -0.230 | 0.034 | 0.006 | 0.037 | -0.202 | 0.038 | 0.003 |
| rs2228603 | T | C | 0.062 | -0.221 | 0.033 | 0.006 | 0.067 | -0.146 | 0.024 | 0.003 |
| rs2229738 | T | C | 0.169 | -0.127 | 0.021 | 0.005 | 0.140 | -0.083 | 0.022 | 0.002 |
| rs75306442 | T | C | 0.187 | -0.105 | 0.020 | 0.003 | 0.193 | -0.044 | 0.017 | 0.001 |

EA: effect allele. NEA: non-effect (other) allele. EAF: effect allele frequency. BETA: beta-coefficient for effect of additional effect allele on inverse-normal transformed total omega-3 fatty acidtty acid levels. $R^2$: proportion of variance in fatty acid explained by each SNP, calculated using the equation $R^2_{snp} = 2\alpha^2$ MAF(1−MAF), where α = the standardized SNP-DHA association and MAF = minor allele frequency.

**Table E6. Characteristics of pooled cohort study participants without spirometry-defined airway obstruction at baseline**

| | OVERALL | CASE | CENSORED |
|---|---|---|---|
| **N PARTICIPANTS** | 11,617 | 1,455 | 10,162 |
| **AGE AT BASELINE, YEARS** | 53.1 (19.2) | 58.4 (16.6) | 52.4 (19.4) |
| **FEMALE SEX, N (%)** | 6,706 (57.7) | 751 (51.6) | 5,955 (58.6) |
| **RACE/ETHNICITY, N (%)** | | | |
| **WHITE** | 7,727 (66.5) | 1,,102 (75.7) | 6,625 (65.2) |
| **BLACK** | 2,555 (22.0) | 184 (12.7) | 2,371 (23.3) |
| **ASIAN** | 511 (4.4) | 86 (5.9) | 425 (4.2) |
| **HISPANIC** | 824 (7.10) | 83 (5.7) | 741 (7.3) |
| **SMOKING STATUS, N (%)** | | | |
| **NEVER** | 6,098 (52.5) | 634 (43.6) | 5,464 (53.8) |
| **FORMER** | 3,794 (32.7) | 563 (38.7) | 3,231 (31.8) |
| **CURRENT** | 1,725 (14.9) | 258 (17.7) | 1,467 (14.4) |
| **SMOKING PACK-YEARS, MEDIAN (IQR)[A]** | 10.5 (24.5) | 17.0 (27.9) | 9.5 (23.4) |
| **SMOKING CIGARETTES PER DAY, MEDIAN (IQR)[B]** | 10 (14) | 15 (13) | 10 (15) |
| **FOLLOW-UP TIME, YEARS, MEDIAN (IQR)** | 5.0 (16.0) | 5.0 (8.1) | 5.0 (17.0) |
| **ALA, %[C]** | 0.2 (0.1) | 0.2 (0.1) | 0.2 (0.1) |
| **EPA, %[C]** | 0.7 (0.6) | 0.7 (0.7) | 0.7 (0.6) |
| **DPA, %[C]** | 0.9 (0.2) | 0.9 (0.2) | 0.9 (0.2) |
| **DHA, %[C]** | 3.3 (1.2) | 3.2 (1.2) | 3.3 (1.2) |

All values reported as mean (SD) unless otherwise noted. Cases defined as participants who transitioned from FEV1/FVC ≥ 0.7 at study baseline to FEV1/FVC < 0.7 at any subsequent spirometry measurement. Participants with FEV1/FVC ≥ 0.7 for all spirometry measurements were censored.

[a] Smoking pack-years calculated among former and current smokers (N = 5,515).

[b] Smoking cigarettes per day calculated among current smokers (N = 1,350).

[c] Percentage of total fatty acids in plasma phospholipids.

**Table E7. Correlations of ALA with EPA, DPA, and DHA**

| | | EPA | | DPA | | DHA | |
|---|---|---|---|---|---|---|---|
| | **N** | **Coeff** | **P-value** | **Coeff** | **P-value** | **Coeff** | **P-value** |
| **OVERALL** | 10,563 | 0.18 | < 0.001 | 0.10 | < 0.001 | 0.03 | < 0.001 |
| **MALE** | 6,839 | 0.19 | < 0.001 | 0.14 | < 0.001 | 0.04 | 0.000 |
| **FEMALE** | 8,224 | 0.17 | < 0.001 | 0.09 | < 0.001 | 0.01 | 0.255 |
| **WHITE** | 10,434 | 0.21 | < 0.001 | 0.10 | < 0.001 | 0.04 | < 0.001 |
| **BLACK** | 3,008 | 0.16 | < 0.001 | 0.08 | < 0.001 | -0.04 | 0.034 |
| **ASIAN** | 641 | 0.04 | 0.266 | 0.08 | 0.033 | -0.04 | 0.324 |
| **HISPANIC** | 980 | 0.11 | 0.001 | 0.06 | 0.055 | -0.09 | 0.004 |
| **NEVER SMOKERS** | 7,169 | 0.16 | < 0.001 | 0.08 | < 0.001 | 0.02 | 0.037 |
| **FORMER SMOKERS** | 5,440 | 0.20 | < 0.001 | 0.12 | < 0.001 | 0.05 | < 0.001 |
| **CURRENT SMOKERS** | 2,454 | 0.20 | < 0.001 | 0.06 | 0.003 | -0.04 | 0.067 |

Coeff: Pearson’s correlation coefficient.

**Table E8. Unadjusted and Benjamini and Hochberg false discovery rate adjusted p-values for three-way interaction terms evaluating effect modification by sex, race/ethnicity, and baseline smoking status**

| | UNADJUSTED P-VALUES | | | FDR ADJUSTED P-VALUES | | |
|---|---|---|---|---|---|---|
| | SEX | SMK | RACE | SEX | SMK | RACE |
| **$FEV_1$ DECLINE[a]** | | | | | | |
| **ALA** | **0.027** | **0.013** | **0.005** | 0.053 | **0.039** | **0.032** |
| **EPA** | 0.174 | 0.065 | **0.005** | 0.261 | 0.112 | 0.059 |
| **DPA** | **0.006** | 0.964 | 0.360 | **0.024** | 0.323 | 0.432 |
| **DHA** | 0.855 | 0.242 | **0.014** | 0.933 | 0.964 | **0.033** |
| **FVC DECLINE[a]** | | | | | | |
| **ALA** | **0.016** | **0.012** | **0.008** | **0.038** | **0.048** | **0.046** |
| **EPA** | 0.814 | **0.002** | **0.014** | 0.888 | **0.022** | **0.041** |
| **DPA** | 0.173 | 0.823 | 0.108 | 0.231 | 0.823 | 0.162 |
| **DHA** | 0.729 | **0.017** | **0.023** | 0.874 | **0.034** | **0.039** |
| **INCIDENT AIRWAY OBSTRUCTION[b]** | | | | | | |
| **ALA** | 0.821 | 0.085 | 0.853 | 0.896 | 1.000 | 0.853 |
| **EPA** | 0.630 | 0.630 | 0.659 | 0.946 | 0.841 | 0.791 |
| **DPA** | 0.204 | 0.214 | 0.188 | 0.816 | 0.641 | 1.000 |
| **DHA** | 0.600 | 0.554 | 0.583 | 1.000 | 1.000 | 1.000 |

Associations with $P < 0.05$ are shown in bold.

FDR: False Discovery Rate. SMK: baseline smoking status.

[a] Type-3 P-values from 3-way interaction terms of sex/baseline smoking status/race with elapsed time and omega-3 fatty acid concentrations in linear mixed effects models of $FEV_1$ or FVC decline.

[b] Type-3 P-values from interaction terms of sex/baseline smoking status/race with omega-3 fatty acid concentrations in cox proportional hazards models of incident airway obstruction ($FEV_1/FVC < 0.7$).

**Table E9. Mendelian Randomization replication analysis results for EPA, DPA, DHA, and total omega-3 fatty acids**

| Fatty acid | Replication GWAS | N SNP | F statistic | MR method | $FEV_1$ (ukb-b-11141) | | FVC (ukb-b-14713) | | $FEV_1/FVC < 0.7$ (ieu-b-106) | |
|---|---|---|---|---|---|---|---|---|---|---|
| | | | | | Beta | [95% CI] | Beta | [95% CI] | OR | [95% CI] |
| EPA | Shin et al. | 2 | NA | Inverse variance weighted | 0.138 | [ 0.023, 0.254] | 0.086 | [-0.024, 0.195] | 0.974 | [0.916, 1.034] |
| DPA | Shin et al. | 3 | NA | MR Egger<br>Weighted median<br>Inverse variance weighted<br>Weighted mode | 0.225<br>0.159<br>0.133<br>0.173 | [-0.054, 0.504]<br>[-0.040, 0.358]<br>[-0.031, 0.297]<br>[ 0.004, 0.341] | 0.134<br>0.079<br>0.060<br>0.103 | [-0.367, 0.635]<br>[-0.105, 0.262]<br>[-0.182, 0.301]<br>[-0.054, 0.260] | 0.961<br>0.966<br>0.962<br>0.969 | [0.763, 1.211]<br>[0.850, 1.098]<br>[0.867, 1.068]<br>[0.903, 1.040] |
| DHA | Kettunen et al. | 5 | 41 | MR Egger<br>Weighted median<br>Inverse variance weighted<br>Weighted mode | 0.065<br>0.033<br>0.017<br>0.037 | [-0.050, 0.180]<br>[ 0.005, 0.061]<br>[-0.023, 0.056]<br>[ 0.007, 0.066] | 0.066<br>0.018<br>0.019<br>0.022 | [-0.039, 0.170]<br>[-0.011, 0.047]<br>[-0.018, 0.055]<br>[-0.009, 0.054] | 1.009<br>0.997<br>1.001<br>0.996 | [0.970, 1.050]<br>[0.984, 1.010]<br>[0.989, 1.014]<br>[0.984, 1.009] |
| | Lemaitre et al. | 2 | 6 | Inverse variance weighted | 0.069 | [-0.005, 0.142] | 0.050 | [-0.087, 0.187] | 0.995 | [0.952, 1.039] |
| | Shin et al. | 2 | NA | Inverse variance weighted | 0.453 | [ 0.125, 0.781] | 0.470 | [ 0.133, 0.807] | 1.048 | [0.820, 1.339] |

| | | | | | | | | | | |
|---|---|---|---|---|---|---|---|---|---|---|
| Total ω-3 | Kettunen et al. | 6 | 44 | MR Egger<br>Weighted median<br>Inverse variance weighted<br>Weighted mode | 0.018<br>0.029<br>0.018<br>0.030 | [-0.039, 0.075]<br>[ 0.006, 0.053]<br>[-0.014, 0.049]<br>[ 0.007, 0.053] | 0.016<br>0.019<br>0.019<br>0.021 | [-0.037, 0.070]<br>[-0.004, 0.042]<br>[-0.011, 0.048]<br>[-0.002, 0.043] | 1.004<br>0.997<br>1.001<br>0.998 | [0.987, 1.021]<br>[0.986, 1.009]<br>[0.991, 1.011]<br>[0.987, 1.009] |

Associations with $p < 0.05$ are shown in bold.

F statistics calculated using the equation $F = (N-K-1)/K \times (R^2_{instrument})/(1-R^2_{instrument})$, where n = sample size in fatty acid GWAS, K = number of SNPs contributing to the genetic instrument and $R^2_{instrument}$=sum of $R^2_{snp}$ across SNPs contributing to the instrument. F statistics could not be calculated for analyses using results from the Shin GWAS because the beta coefficients could not be converted to the required units for calculation of $R^2$.

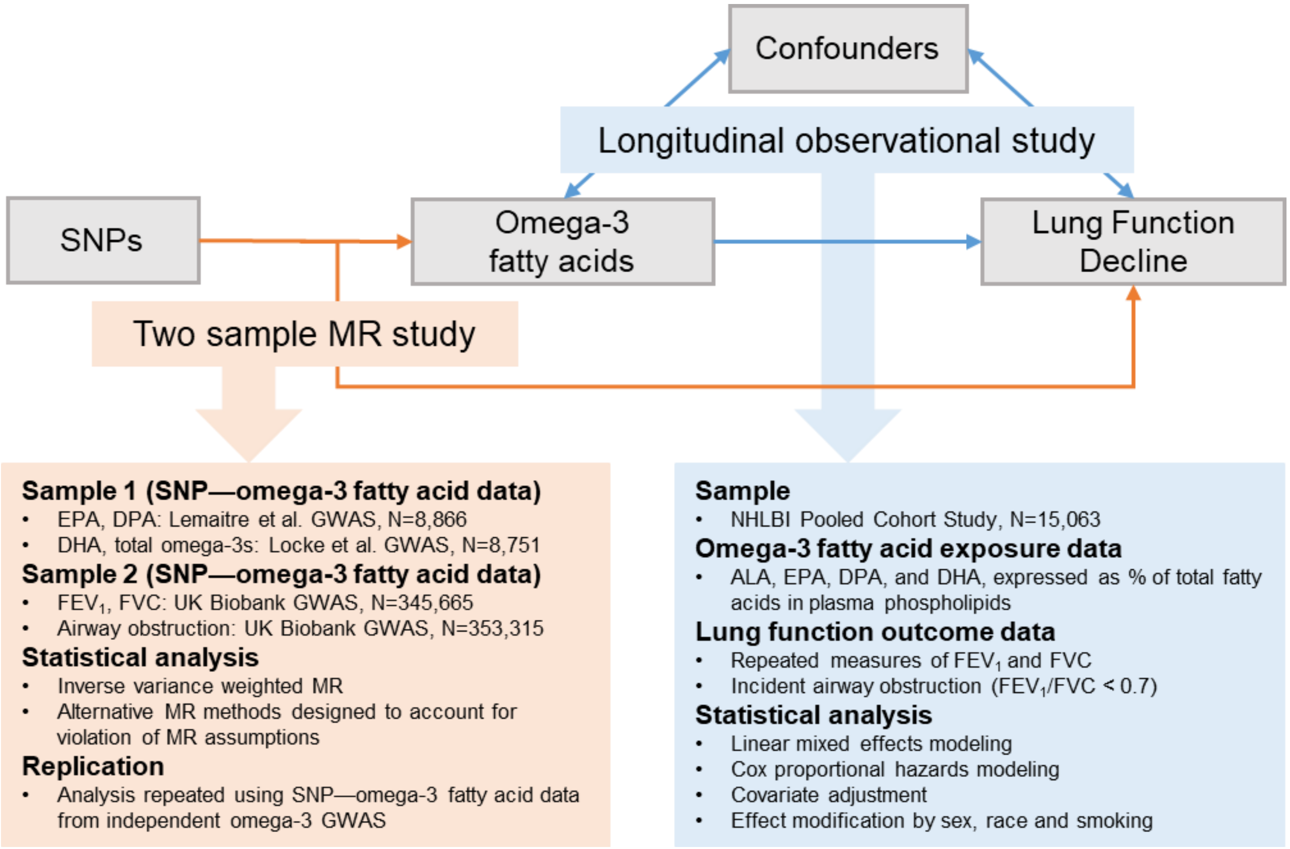


**Figure E1. Overview of the study design.** SNPs: Single Nucleotide Polymorphisms. Diagram showing the two complementary study designs used in this study. The longitudinal cohort study was performed to estimate the associations of omega-3 fatty acids with decline in $FEV_1$ and FVC and the development of spirometry-defined airway obstruction. The two-sample MR study was performed to estimate the associations of genetically predicted omega-3 fatty acids with $FEV_1$, FVC, and odds of spirometry-defined airway obstruction.

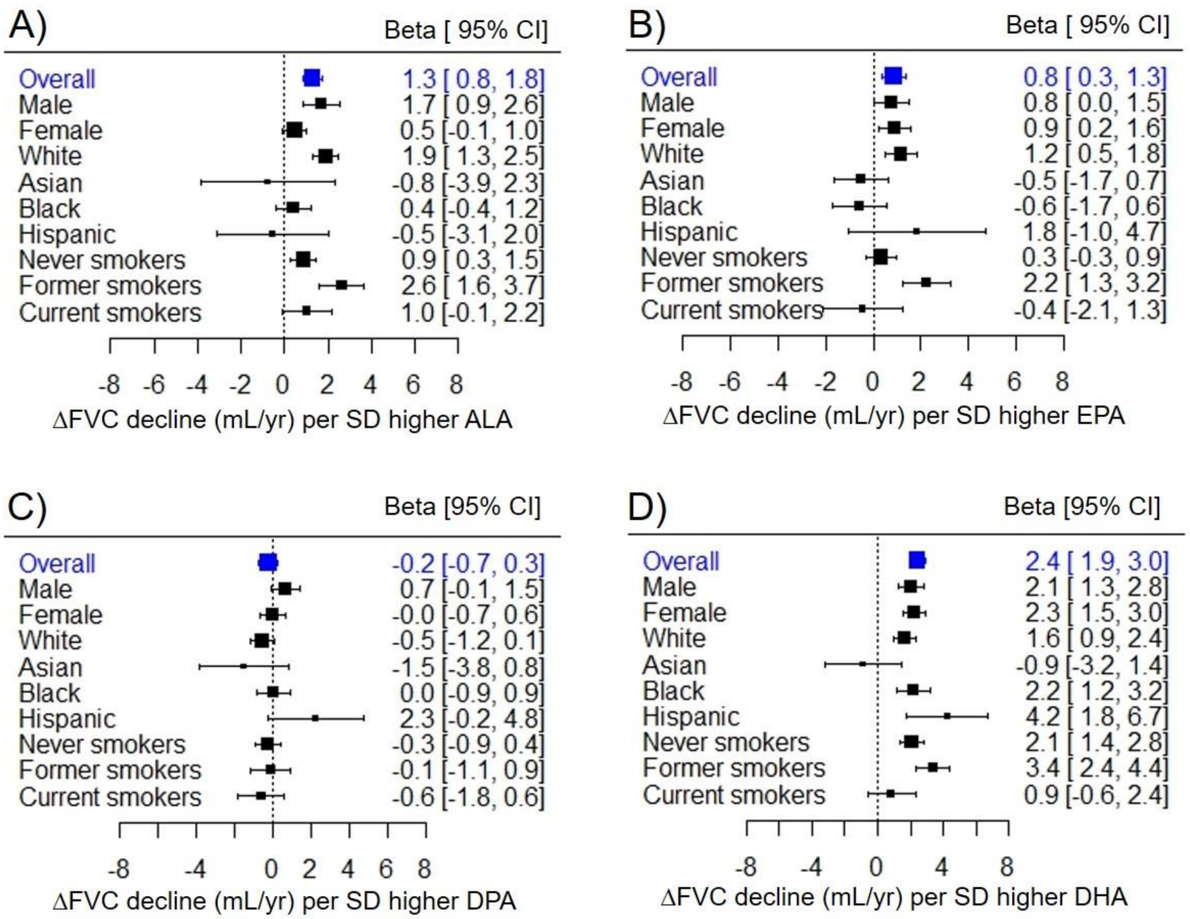


**Figure E2. Effect modification of omega-3 fatty acid—FVC decline associations by sex, race and/or baseline smoking status.** Sex, race/ethnicity, and baseline smoking status differences in associations of A) ALA, B) EPA, C) DPA, and D) DHA with FVC decline. Estimates (Betas) and 95% confidence intervals (CIs) for each group were derived from three-way interaction terms and covariances. All estimates are presented per standard deviation (SD) higher fatty acid.

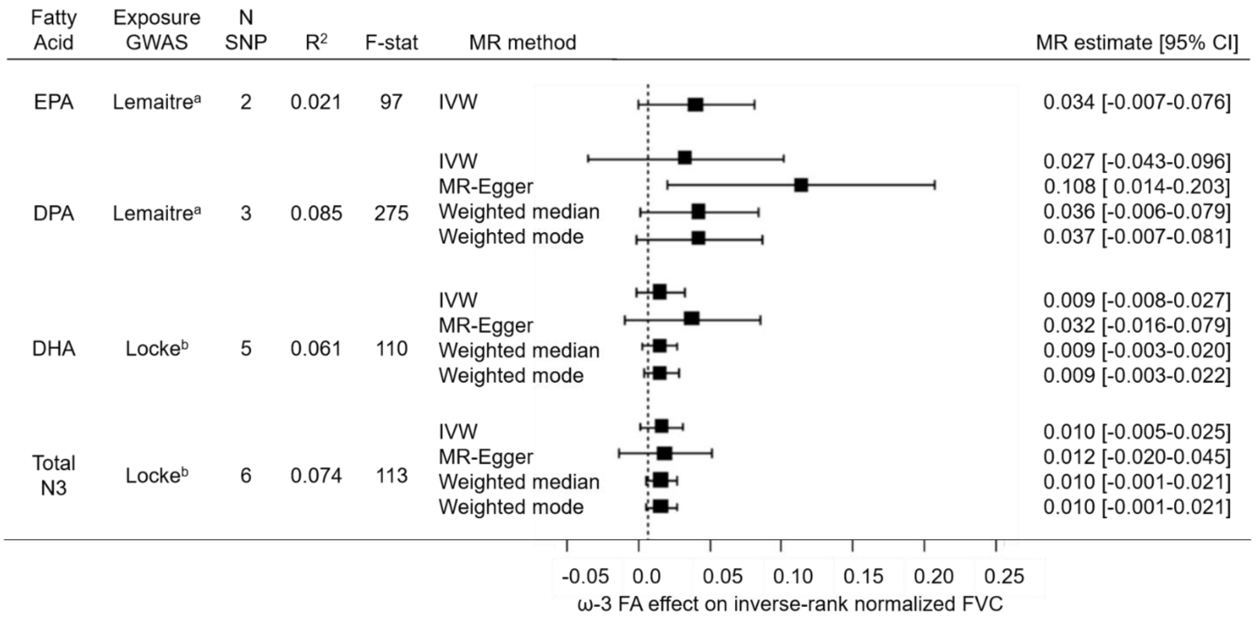


**Figure E3. Forest plot of two-sample Mendelian Randomization estimates from primary analysis of EPA, DPA, DHA, and total omega-3 fatty acids on FVC.** Forest plot showing MR estimates and 95% confidence intervals (CIs) for associations of genetically predicted omega-3 fatty acids with FVC using different MR methods.

N SNP: number of SNPs contributing to the omega-3 fatty acid genetic instrument.

Exposure GWAS: GWAS that SNP—omega-3 fatty acid association data were extracted from.

$R^2$: proportion of omega-3 fatty acid variance explained by the genetic instrument.

F-stat: F statistic, a measure of genetic instrument strength where the MR standard for validity is F > 10. IVW: Inverse variance weighted MR.

[a] Lemaitre et al. GWAS fatty acid units in % of total fatty acid composition

[b] Locke et al. GWAS fatty acid units in standardized fatty acid concentration (mmol/L)

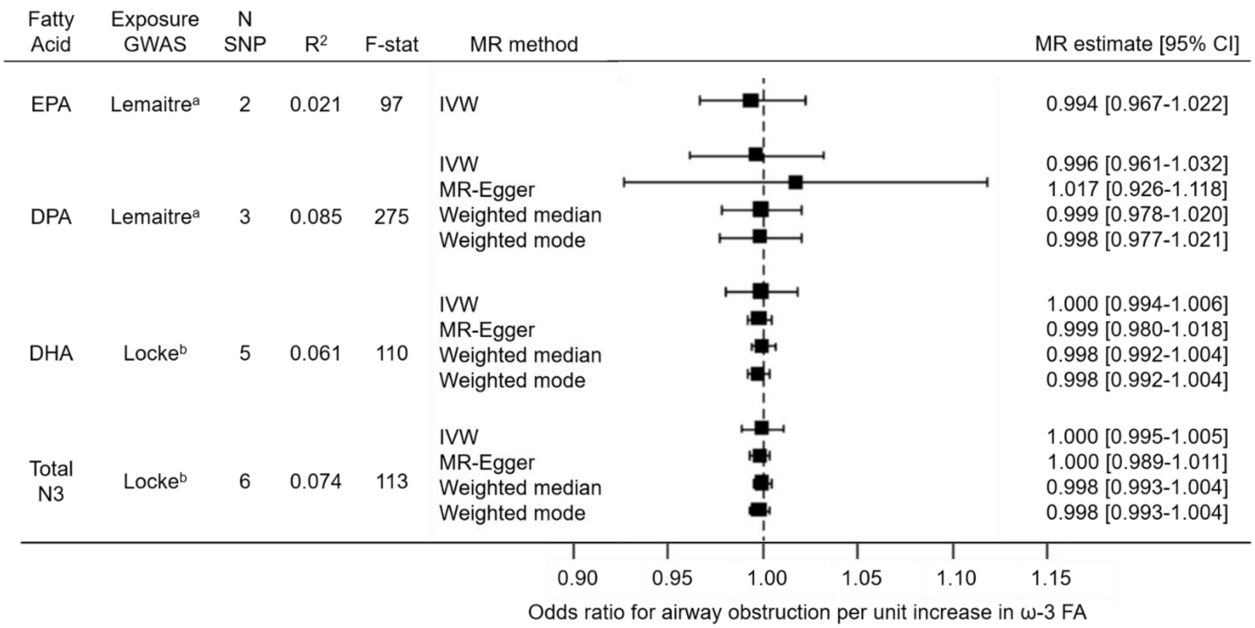


**Figure E4. Forest plot of two-sample Mendelian Randomization estimates from primary analysis of EPA, DPA, DHA and total omega-3 fatty acids on airway obstruction.** Forest plot showing MR estimates and 95% confidence intervals (CIs) for associations of genetically predicted omega-3 fatty acids with odds or spirometry-defined airway obstruction using different MR methods.

N SNP: number of SNPs contributing to the omega-3 fatty acid genetic instrument.

Exposure GWAS: GWAS that SNP—omega-3 fatty acid association data were extracted from.

$R^2$: proportion of omega-3 fatty acid variance explained by the genetic instrument.

F-stat: F statistic, a measure of genetic instrument strength where the MR standard for validity is F>10. IVW: Inverse variance weighted MR.

[a] Lemaitre et al. GWAS fatty acid units in % of total fatty acid composition

[b] Locke et al. GWAS fatty acid units in standardized fatty acid concentration (mmol/L)
