## Supplementary material for "Investigating associations of omega-3 fatty acids, lung function decline, and airway obstruction": STROBE Checklists

**STROBE checklist for longitudinal study**

| Recommendation | Page Number |
| --- | --- |
| **Title and abstract** | |
| *(a) Indicate the study’s design with a commonly used term in the title or the abstract*  *(b) Provide in the abstract an informative and balanced summary of what was done and what was found* | a) Page 1 – abstract: methods  b) Title page – study summary |
| **Introduction** | |
| Background  *Explain the scientific background and rationale for the investigation being reported* | Pages 2-3 |
| Objectives  *State specific objectives, including any prespecified hypotheses* | Page 3 |
| **Methods** | |
| Study design  *Present key elements of study design early in the paper* | Page 3 |
| Setting  *Describe the setting, locations, and relevant dates, including periods of recruitment, exposure, follow-up, and data collection* | Pages 3-4; online data supplement pages Table E1 |
| Participants  *(a) Give the eligibility criteria, and the sources and methods of selection of participants. Describe methods of follow-up*  *(b) For matched studies, give matching criteria and number of exposed and unexposed* | (a) Page 3; online data supplement pages Table E1  (b) NA |
| Variables  *Clearly define all outcomes, exposures, predictors, potential confounders, and effect modifiers. Give diagnostic criteria, if applicable* | Pages 3-4 |
| Data sources/management  *For each variable of interest, give sources of data and details of methods of assessment (measurement). Describe comparability of assessment methods if there is more than one group* | Pages 3-4; online data supplement pages Table E1 |
| Bias  *Describe any efforts to address potential sources of bias* | Pages 4-5 |
| Study size  *Explain how the study size was arrived at* | Pages 3-4 |
| Quantitative variables  *Explain how quantitative variables were handled in the analyses. If applicable, describe which groupings were chosen and why* | Page 4; online data supplement page 2 |
| Statistical methods  *(a) Describe all statistical methods, including those used to control for confounding*  *(b) Describe any methods used to examine subgroups and interactions*  *(c) Explain how missing data were addressed*  *(d) If applicable, explain how loss to follow-up was addressed*  *(e) Describe any sensitivity analyses* | (a) Page 4; online data supplement page 2  (b) Page 5  (c) NA  (d) Online data supplement page 2 (censoring for cox proportional hazards modelling)  (e) Online data supplement page 2 |
| **Results** | |
| Participants  *(a) Report numbers of individuals at each stage of study—eg numbers potentially eligible, examined for eligibility, confirmed eligible, included in the study, completing follow-up, and analysed*  *(b) Give reasons for non-participation at each stage*  *(c) Consider use of a flow diagram* | (a) Page 4; online data supplement page 2  (b) Page 5  (c) NA  (d) Online data supplement page 2 (censoring for cox proportional hazards modelling)  (e) Online data supplement page 2 |
| Descriptive data  *(a) Give characteristics of study participants (eg demographic, clinical, social) and information on exposures and potential confounders*  *(b) Indicate number of participants with missing data for each variable of interest*  *(c) Summarise follow-up time (eg, average and total amount)* | (a) Page 6; Table 1, online data supplement Table E6  (b) Table 1  (c) Page 6; page 8; Table 1; online data supplement Table E6 |
| Outcome data  *Report numbers of outcome events or summary measures over time* | Page 8; Table 1; online data supplement Table E6 |
| Main results  *(a) Give unadjusted estimates and, if applicable, confounder-adjusted estimates and their precision (eg, 95% confidence interval). Make clear which confounders were adjusted for and why they were included*  *(b) Report category boundaries when continuous variables were categorized*  *(c) If relevant, consider translating estimates of relative risk into absolute risk for a meaningful time period* | (a) Page 7; Table 2; Figure 2  (b) NA  (c) NR |
| Other analyses  *Report other analyses done—eg analyses of subgroups and interactions, and sensitivity analyses* | Pages 7-8; Figure 1; online data supplement Table E8 |
| **Discussion** | |
| Key results  *Summarize key results with reference to study objectives.* | Pages 10-12 |
| Limitations  *Discuss limitations of the study, taking into account sources of potential bias or imprecision. Discuss both direction and magnitude of any potential bias* | Pages 13-14 |
| Interpretation  *Give a cautious overall interpretation of results considering objectives, limitations, multiplicity of analyses, results from similar studies, and other relevant evidence* | Page 14 |
| Generalizability  *Discuss the generalisability (external validity) of the study results* | Pages 11-12 |
| **Other information** | |
| Funding  *Give the source of funding and the role of the funders for the present study and, if applicable, for the original study on which the present article is based* | Title page |

**STROBE-MR checklist for Mendelian Randomization Study**

| Recommendation | Page Number |
| --- | --- |
| **Title and abstract** | |
| *Indicate Mendelian randomization as the study’s design in the title and/or the abstract.* | Page 1 – abstract: methods |
| **Introduction** | |
| Background  *Explain the scientific background and rationale for the reported study. Is causality between exposure and outcome plausible? Justify why MR is a helpful method to address the study question.* | Pages 2-3 |
| Objectives  *State specific objectives clearly, including pre-specified causal hypotheses (if any).* | Page 3 |
| **Methods** | |
| Study design and data sources  *a) Describe the study design and the underlying population from which it was drawn.*  *Describe also the setting, locations, and relevant dates, including periods of recruitment,*  *exposure, follow-up, and data collection, if available.*  *b) Give the eligibility criteria, and the sources and methods of selection of participants.*  *c) Explain how the analyzed sample size was arrived at.*  *d) Describe measurement, quality and selection of genetic variants.*  *e) For each exposure, outcome and other relevant variables, describe methods of assessment*  *and, in the case of diseases, the diagnostic criteria used.*  *f) Provide details of ethics committee approval and participant informed consent, if relevant.* | a, b, c) Page 3; online data supplement pages 3-4; online data supplement figure E1  d, e) Page 5; online data supplement pages 3-4  f) NA |
| Assumptions  *Explicitly state assumptions for the main analysis (e.g. relevance, exclusion, independence, homogeneity) as well assumptions for any additional or sensitivity analysis.* | Page 6 |
| Statistical methods: main analysis  a) Describe how quantitative variables were handled in the analyses (i.e., scale, units,  model).  b) Describe the process for identifying genetic variants and weights to be included in the  analyses (i.e, independence and model). Consider a flow diagram.  c) Describe the MR estimator, e.g. two-stage least squares, Wald ratio, and related statistics.  Detail the included covariates and, in case of two-sample MR, whether the same  covariate set was used for adjustment in the two samples.  d) Explain how missing data were addressed.  e) If applicable, say how multiple testing was dealt with. | a) Figure 3; online data supplement Figures E3-E4  b) Page 6; online data supplement paged 3-4  c) Page 6  d) NA  e) NA |
| Assessment of assumptions  *Describe any methods used to assess the assumptions or justify their validity.* | Page 6 |
| Sensitivity analyses  *Describe any sensitivity analyses or additional analyses performed.* | Page 5; online data supplement pages 3-4 |
| Software and pre-registration  a) Name statistical software and package(s), including version and settings used.  b) State whether the study protocol and details were pre-registered (as well as when and where). | a) Page 6  b) NA |
| **Results** | |
| Descriptive data  *a) Report the numbers of individuals at each stage of included studies and reasons for exclusion. Consider use of a flow-diagram.*  *b) Report summary statistics for phenotypic exposure(s), outcome(s) and other relevant variables (e.g. means, standard deviations, proportions).*  *c) If the data sources include meta-analyses of previous studies, provide the number of studies, their reported ancestry, if available, and assessments of heterogeneity across these studies. Consider using a supplementary table for each data source.*  *d) For two-sample Mendelian randomization:*  *i. Provide information on the similarity of the genetic variant-exposure associations between the exposure and outcome samples.*  *ii. Provide information on extent of sample overlap between the exposure and outcome data sources.* | a) Online data supplement pages 3-4; online data supplement Figure E1 b) NR c) Online data supplement pages 3-4 d) Online data supplement pages 3-4 |
| Main results  *a) Report the associations between genetic variant and exposure, and between genetic variant and outcome, preferably on an interpretable scale (e.g. comparing 25th and 75^th^ percentile of allele count or genetic risk score, if individual-level data available).*  *b) Report causal effect estimate between exposure and outcome, and the measures of uncertainty from the MR analysis. Use an intuitive scale, such as odds ratio, or relative risk, per standard deviation difference.*  *c) If relevant, consider translating estimates of relative risk into absolute risk for ameaningful time-period.*  *d) Consider any plots to visualize results (e.g. forest plot, scatterplot of associations between*  *genetic variants and outcome versus between genetic variants and exposure).* | a) Online data supplement Tables E2-E5 b) Figure 3; online data supplement Figures E3-E4; online data supplement Table E9 c) NA d) Figure 3; online data supplement Figures E3-E4 |
| Assessment of assumptions  *a) Assess the validity of the assumptions.*  *b) Report any additional statistics (e.g., assessments of heterogeneity, such as I2, Q statistic).* | a) Pages 9-10  b) NR |
| Sensitivity and additional analyses  *a) Use sensitivity analyses to assess the robustness of the main results to violations of the assumptions.*  *b) Report results from other sensitivity analyses (e.g., replication study with different dataset, analyses of subgroups, validation of instrument(s), simulations, etc.).*  *c) Report any assessment of direction of causality (e.g., bidirectional MR).*  *d) When relevant, report and compare with estimates from non-MR analyses.*  *e) Consider any additional plots to visualize results (e.g., leave-one-out analyses).* | a) Pages 9-10  b) Page 10; online data supplement Table E9  c) NA  d) Pages 10-11  e) NA |
| Key results  *Summarize key results with reference to study objectives.* | Pages 10-12 |
| Limitations  *Discuss limitations of the study, taking into account the validity of the MR assumptions, other sources of potential bias, and imprecision. Discuss both direction and magnitude of any potential*  *bias, and any efforts to address them.* | Page 10; pages 13-14 |
| Interpretation  *a) Give a cautious overall interpretation of results considering objectives and limitations. Compare with results from other relevant studies.*  *b) Discuss underlying biological mechanisms that could be modelled by using the genetic variants to assess the relationship between the exposure and the outcome.*  *c) Discuss whether the results have clinical or policy relevance, and whether interventions could have the same size effect.* | a) Pages 11-13; page 14  b) Page 11  c) Page 14 |
| Generalizability  *Discuss the generalizability of the study results (a) to other populations (i.e. external validity),*  *(b) across other exposure periods/timings, and (c) across other levels of exposure.* | a) Pages 11-12  b and c) NR |
| Funding | Title page |
